# A Deep Learning Method to Detect Opioid Prescription and Opioid Use Disorder from Electronic Health Records

**DOI:** 10.1101/2021.09.13.21263524

**Authors:** Aditya Kashyap, Chris Callison-Burch, Mary Regina Boland

**Affiliations:** Department of Computer Science, University of Pennsylvania; Department of Biostatistics, Epidemiology and Informatics, Perelman School of Medicine, University of Pennsylvania; Institute for Biomedical Informatics, University of Pennsylvania; Center for Excellence in Environmental Toxicology, University of Pennsylvania; Department of Biomedical and Health Informatics, Children’s Hospital of Philadelphia

**Keywords:** opioid, machine learning, electronic health records, data mining, natural language processing

## Abstract

**Objective:** As the opioid epidemic continues across the United States, methods are needed to accurately and quickly identify patients at risk for opioid use disorder (OUD). The purpose of this study is to develop two predictive algorithms: one to predict opioid prescription and one to predict OUD.

**Materials and Methods:** We developed an informatics algorithm that trains two deep learning models over patient EHRs using the MIMIC-III database. We utilize both the structured and unstructured parts of the EHR and show that it is possible to predict both of these challenging outcomes.

**Results:** Our deep learning models incorporate both structured and unstructured data elements from the EHRs to predict opioid prescription with an F1-score of 0.88 ± 0.003 and an AUC-ROC of 0.93 ± 0.002. We also constructed a model to predict OUD diagnosis achieving an F1-score of 0.82 ± 0.05 and AUC-ROC of 0.94 ± 0.008.

**Discussion:** Our model for OUD prediction outperformed prior algorithms for specificity, F1 score and AUC-ROC while achieving equivalent sensitivity. This demonstrates the importance of a.) deep learning approaches in predicting OUD and b.) incorporating both structured and unstructured data for this prediction task. No prediction models for opioid prescription as an outcome were found in the literature and therefore this represents an important contribution of our work as opioid prescriptions are more common than OUDs.

**Conclusion:** Algorithms such as those described in this paper will become increasingly important to understand the drivers underlying this national epidemic.

## 1. BACKGROUND AND SIGNIFICANCE

### 1.1 Background of the Opioid Epidemic in the USA

In 2017, 70,200 drug overdose deaths occurred in the USA with the sharpest increases observed among fentanyl and fentanyl analogs [1]. The number of opioid prescriptions was 81.3 per 100 people, peaking in 2012 [2]. Even with efforts to mitigate the steady rise in opioid prescribing, there were still 58.5 prescriptions per 100 people in 2017 [2]. More than 17% of Americans had a prescription opioid filled in 2017 [2]. The perceived overprescribing of opioids followed by drug-induced overdoses has been termed the ‘opioid epidemic’ [3]. Methods for identifying the causes of this epidemic and identifying high-risk individuals are needed to guide clinicians’ decision making [4].

The opioid epidemic is currently on the rise, with almost 50,000 people in USA having lost their lives due to an opioid related overdose in 2019 alone [5]. This epidemic not only increases the overall national mortality rate, but also creates an “economic burden” estimated at around $78.5 billion a year [5]. The opioid epidemic only worsened during the 2020 COVID-19 pandemic with one study reporting that opioid overdoses were 29% higher during the 2020 COVID-19 pandemic than before[6]. The reason for this increase is multi-faceted with contributing factors including the suspension of 12 Step programs due to the requirements of social distancing that forced the suspensions of their meetings. In addition, the economic distress of the pandemic may have exacerbated the effects of the opioid epidemic on already marginalized communities. Therefore, the need to prevent and detect opioid related problems has never been greater.

### 1.2 Use of Machine Learning Models with Electronic Health Records data

Electronic Health Records (EHRs) are increasingly being used to perform retrospective analyses to inform clinical care - a critical component of the Learning Health System [7, 8]. Retrospective analysis of EHRs can address some of the major challenges of the opioid epidemic by identifying high-risk patient groups and thereby provide much needed information to clinicians. EHRs can also be used to tease apart the multifactorial issues involved with both pain assessment and management, and prescribing behavior involved in the opioid epidemic [4].

As EHRs contain multimodal patient information like medical history, diagnoses, medications, treatment plans, immunization dates, allergies, radiology images, and laboratory and test results, techniques from Computer Vision (CV), Natural Language Processing (NLP), Speech Recognition and the traditional Machine Learning (ML) and Deep Learning fields need to be applied when analyzing them. Researchers have built models on EHRs to study several different medical outcomes in the past, including Alzheimer’s disease [9, 10], osteoarthritis[11], breast nodules and lesions [12], diabetic retinopathy [13], skin cancer [14], heart failure and chronic obstructive pulmonary disease [15], and many others. [16-24]. More recently, due to the pandemic, a lot of research has focused on building models that predict risk of COVID-19 in the general population [25-27], detect COVID-19 in patients with suspected infections [28-30] and provide prognosis for patients with COVID-19 diagnosis [31-34].

### 1.3 Use of Machine Learning Models to predict Opioid Dependence or Use Disorder

A considerable amount of work has also been done in studying opioid dependence in the clinical setting. Previously, researchers searched the OVID database for work related to designing opioid abuse predictive models, and reviewed 7 papers and 9 models with 75 distinct variables [35]. They found that a majority of the papers depended on the presence of diagnostic codes from the International Classification of Diseases (ICD) version 9 (ICD-9) codes to define opioid abuse. They conclude that age and gender were the most consistent demographic variables in predicting opioid abuse (commonly referred to today as Opioid Use Disorder or OUD but in diagnostic codes still reported as opioid abuse) and that utilizing unstructured data in future research may lead to improved model performances. Authors in [36] trained a Random Forest Classifier on patient EHRs to predict opioid dependence and found that opioid dependent patients had significantly higher White Blood Cell (WBC) markers and respiratory disturbances. They also found opioid dependent patients to be commonly malnourished and suffer from psychiatric disorders. Researchers trained an LSTM [38] model with attention to predict opioid overdose risk for patients prescribed opioids from EHRs [37]. They found the most important features for identifying patients at risk of overdose to be the mention of opioids, pain scale score, anti-propulsives, general anesthetics and alcohol use among others. Other researchers found that there was an alarming rate of chronic pain conditions occurring before the onset of OUD and that the associated severe mental health and physical health conditions require better models for assessment [39]. Another group of researchers used NLP to process clinical text to identify opioid related overdoses [40]. They used MediClass [41] to first extract medical concepts from the text, following which they used a manually constructed “rule tree” to classify opioid related overdoses [40].

### 1.4 Development of Deep Learning and NLP

With recent breakthroughs in Deep Learning and NLP, there are several opportunities to build clinical decision models that are more accurate and are able to leverage multimodal data. Transformers [42] are model architectures that are based solely on attention mechanisms that have several advantages over LSTMs. Due to self-attention, they are able to model long range dependencies better. Unlike LSTMs, which are sequential and suffer from the vanishing gradient problem over long sequences, Transformers are able to analyze long input sequences effectively making them an attractive component in recent Deep Learning models. The introduction of Transformers has also led to breakthroughs in NLP. BERT [43] is a language representation model essentially containing stacked multi-layer bidirectional transformers that has enabled transfer learning for NLP tasks and resulted in state-of-the-art performances for a surprising number of tasks. BERT was trained on English Wikipedia and the BooksCorpus [44], and therefore does not transfer well across to the medical/clinical domain. Some informatics approaches solve this problem by pretraining BERT on PubMed abstracts and Pubmed Central (PMC) full-text articles [45]. Researchers built on this work by also including clinical notes from MIMIC-III v1.4 database [46] in the pre-training stage, resulting in ClinicalBERT which achieves state-of-the-art results across several clinical NLP tasks [47].

### 1.5 Purpose of this study

The purpose of this study is to develop a method that incorporates deep learning methods to predict both opioid prescription and OUD using both structured and unstructured EHR data. This will serve as a proof-of-concept towards building point of care algorithms to predict opioid prescribing and OUD to address the opioid epidemic.

## 2. MATERIALS AND METHODS

In this paper, we build two models, one that predicts opioid prescription and the other that predicts OUD diagnosis. Details about the data collection pipeline, along with descriptions of the two models are presented in the following sections.

### 2.1 Dataset

For our experiments, we use the MIMIC-III dataset [46] containing de-identified health data associated with 53,423 distinct Intensive Care Unit (ICU) admissions. The information includes procedures carried out, problems diagnosed, medications prescribed, and clinical notes written during a patient’s treatment by healthcare professionals. Most of this data contains an associated timestamp that we use in our analysis. The characteristics of the patient population are given in **Table 1**. A patient’s age was calculated as the time interval between their Admission Date and Date of Birth, and the outliers as a result of the deidentification process described in [46] were removed (patients > 300 years of age). The remaining data was used in constructing models as described in the following sections.

**Table 1:** Population characteristics of the patients who were prescribed opioids (Pos) and who were not (Neg), and who were OUD patients (Pos) and who were not OUD (Neg)

| Static Features |  | Opioid Prescribed Patients (%) |  | OUD Patients (%) |  |
| --- | --- | --- | --- | --- | --- |
|  |  | Pos | Neg | Pos | Neg |
| Admission Location | CLINIC REFERRAL/PREMATURE | 19.70 | 20.40 | 26.21 | 22.80 |
|  | ADMISSION_LOCATION_EMERGENCY ROOM ADMIT | 40.49 | 33.00 | 52.94 | 36.56 |
|  | PHYS REFERRAL/NORMAL DELI | 19.79 | 18.06 | 5.76 | 19.39 |
|  | TRANSFER FROM HOSP/EXTRAM | 17.00 | 9.58 | 14.41 | 14.94 |
|  | UNKNOWN | 3.00 | 18.93 | 0.65 | 6.29 |
| Admission Type | ELECTIVE | 16.33 | 3.34 | 3.40 | 11.27 |
|  | EMERGENCY | 78.77 | 54.53 | 95.41 | 71.55 |
|  | NEWBORN | 0.68 | 21.62 | 0 | 9.82 |
|  | UNKNOWN | 2.43 | 17.67 | 0 | 5.24 |
|  | URGENT | 1.77 | 2.82 | 1.17 | 2.09 |
| Age Group | 0 – 18 | 0.93 | 22.56 | 0.13 | 10.61 |
|  | 18 – 25 | 2.52 | 1.72 | 7.73 | 1.57 |
|  | 25 – 35 | 3.95 | 2.68 | 19.00 | 3.80 |
|  | 35 – 50 | 13.65 | 7.94 | 36.69 | 11.00 |
|  | 50 – 65 | 28.11 | 14.38 | 33.15 | 24.11 |
|  | 65 + | 48.37 | 33.02 | 3.27 | 43.64 |
|  | UNKNOWN | 2.43 | 17.67 | 0 | 5.24 |
| Ethnicity | ASIAN | 1.40 | 2.48 | 0.13 | 1.44 |
|  | BLACK/AFRICAN AMERICAN | 7.86 | 9.17 | 10.61 | 8.25 |
|  | HISPANIC OR LATINO | 2.60 | 2.60 | 3.67 | 2.35 |
|  | OTHER | 2.18 | 2.30 | 3.54 | 2.23 |
|  | PATIENT DECLINED TO ANSWER | 0.98 | 0.68 | 0.39 | 1.04 |
|  | UNKNOWN | 14.64 | 28.92 | 11.27 | 17.03 |
|  | WHITE | 70.31 | 53.81 | 70.38 | 67.62 |
| Gender | FEMALE | 42.37 | 37.15 | 34.47 | 43.12 |
|  | MALE | 55.19 | 45.17 | 65.53 | 51.63 |
|  | UNKNOWN | 2.43 | 17.67 | 0 | 5.24 |
| Insurance | Government | 2.60 | 2.86 | 7.86 | 2.22 |
|  | Medicaid | 8.80 | 8.98 | 41.80 | 8.25 |
|  | Medicare | 52.07 | 35.59 | 28.83 | 46.26 |
|  | Private | 33.16 | 33.93 | 18.21 | 36.69 |
|  | Self-Pay | 0.92 | 0.93 | 3.27 | 1.31 |
|  | UNKNOWN | 2.43 | 17.67 | 0 | 5.24 |
| Marital Status | DIVORCED | 6.33 | 3.26 | 9.04 | 6.42 |
|  | MARRIED | 47.51 | 26.26 | 16.64 | 41.80 |
|  | SEPARATED | 1.05 | 0.61 | 2.62 | 0.92 |
|  | SINGLE | 24.04 | 16.64 | 58.32 | 19.79 |
|  | UNKNOWN | 7.91 | 43.35 | 9.30 | 19.39 |
|  | WIDOWED | 13.13 | 9.85 | 4.06 | 11.66 |
| Religion | CATHOLIC | 35.98 | 26.31 | 39.18 | 34.07 |
|  | EPISCOPALIAN | 1.31 | 0.96 | 0.78 | 1.17 |
|  | JEWISH | 8.77 | 8.09 | 2.22 | 7.34 |
|  | OTHER | 4.28 | 4.00 | 3.54 | 3.54 |
|  | PROTESTANT QUAKER | 12.23 | 9.38 | 10.09 | 10.61 |
|  | UNKNOWN | 37.40 | 51.23 | 44.16 | 43.25 |

#### 2.1.1 Obtaining a Dataset for the Opioid Prescription Prediction Model

We constructed a binary classification model that can predict whether a patient is likely to be prescribed an opioid using their EHR data. Using a list of 30 opioids (Supplemental Table 1), we parse through the MIMIC-III dataset (to be precise, the Prescriptions and InputEvents sections of the MIMIC-III data) to identify all Hospital Admission IDs (HADM_IDs) where patients have been prescribed any of these drugs. These are taken as positive examples while the remaining set of HADM_IDs (for which patients had not been prescribed an opioid) are considered as negative examples.

For each HADM_ID, we retrieve 3 types of data (if available for that patient during that admission) from MIMIC-III:

1. **Structured static data:** This data includes features of a patient that remain constant during the course of a single admission. Example features include patients’ gender, ethnicity, marital status, religion, and so forth. Certain features are then combined (for example, variants of the ethnicity Hispanic/Latino like Central American, Columbian, Cuban, Dominican, were all grouped together). We grouped features up a level using the hierarchy provided within the MIMIC-III dataset. Finally, any features that occur in less than 1% of the MIMIC-III dataset were removed. We introduced an additional “UNKNOWN” feature for each of the 8 feature categories (**Table 1)** to account for missing data.
2. **Structured Time Varying data:** This data includes information about a patient that typically changes over the course of an admission. These include events such as procedures and lab tests performed, and input/output events. We do not use the value of these events, but rather just record the presence or absence of them. For example, if a patient’s blood pressure is measured, we do not record the numeric value, but rather that it was measured.
3. **Unstructured Clinical Notes** This data includes notes written by healthcare workers while treating patients. The process of obtaining input features for the positive and negative examples differs slightly. For each patient belonging to the positive set (i.e., received an opioid prescription), we initially identify the date and time at which they were first prescribed an opioid. The input features that we consider include all information recorded about the patient in the MIMIC-III EHR before that point in time. This is illustrated in **Figure 1**. We remove any mentions of opioids in the clinical notes and the input events of the positive examples to ensure that information leakage does not occur in our feature space. This is done in order to prevent the prediction task from becoming trivial by allowing the model to look for mentions of opioids (some doctors mention in the notes that they will prescribe an opioid in the future).

**Figure 1:**
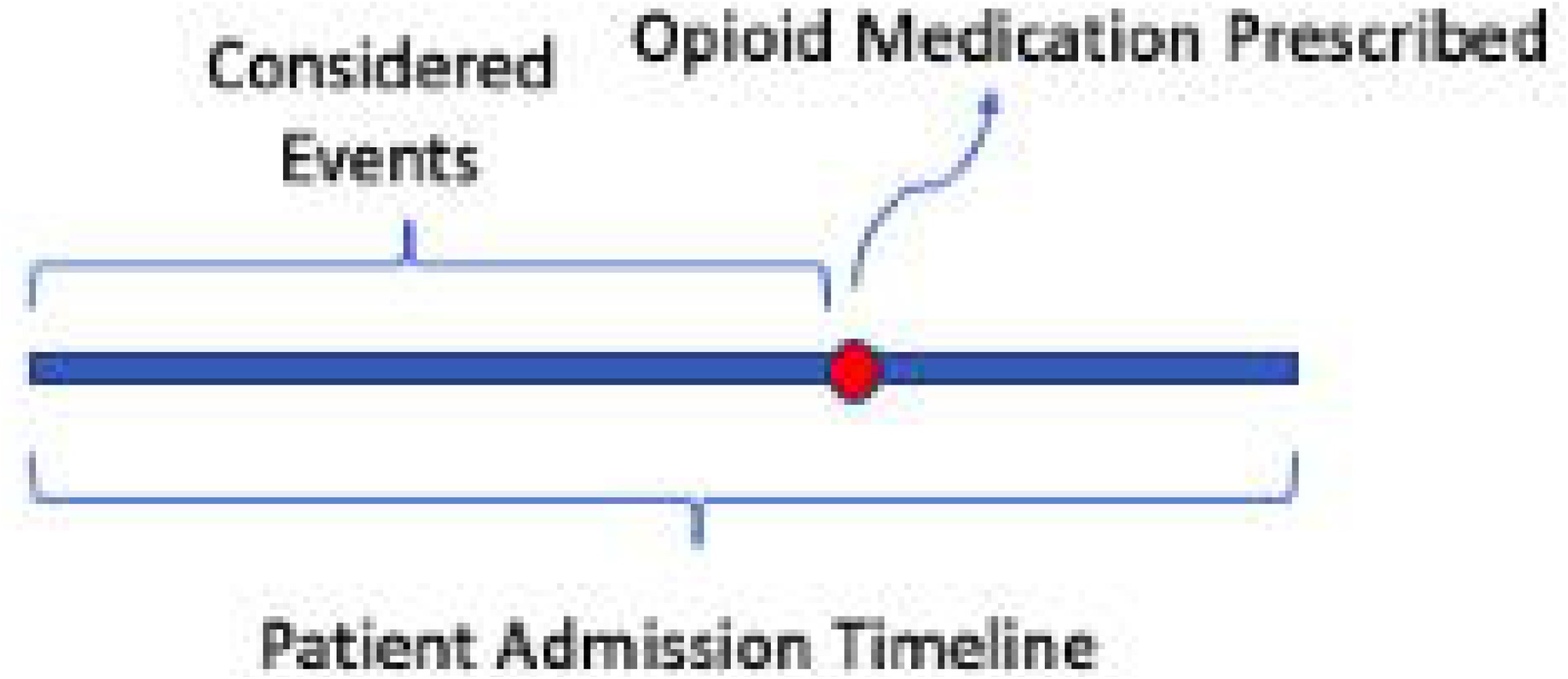
An illustration of a patient admission timeline

Obtaining features for the negative examples is similar. As we do not have a point in time when the opioid was prescribed for these patients, we consider all the information contained about that patient admission in the MIMIC-III EHR. We ensure that only one hospital admission per patient is present across both the positive and negative example sets. This allows the opioid prescription prediction to occur within the course of a single hospital admission timeframe as this was deemed to be clinically more informative.

#### 2.1.2 Obtaining a Dataset for the Opioid Use Disorder (OUD) Prediction Model

Similar to the prescription prediction task, we aim to build a binary classification model that takes as input patient EHR data from MIMIC-III and predicts how likely they are to be diagnosed with OUD. We identify a set of 12 ICD-9 diagnoses codes (Supplemental Table 2) that are given to patients who were either currently showing or who have a history of OUD. The HADM_IDs of patients in MIMIC-III who have been diagnosed with any of these codes were taken as positive examples and the remaining set of HADM_IDs were taken as negative examples. Similar to the prescription prediction task, all the patient admission structured static data, structured time varying data and unstructured clinical notes were used as input to the model.

### 2.2 Model Construction and Training

The binary classification model architectures for both prediction tasks are equivalent and are shown in **Figure 3**. There are 4 components of the model, one for each datatype and a final one that collectively processes information from the first three components. The embeddings computed for each of the 3 datatypes were equal in size (1 × 50) to ensure an equal representation of information across the different modalities. The 4 components of the model are described below:

**Figure 2:**
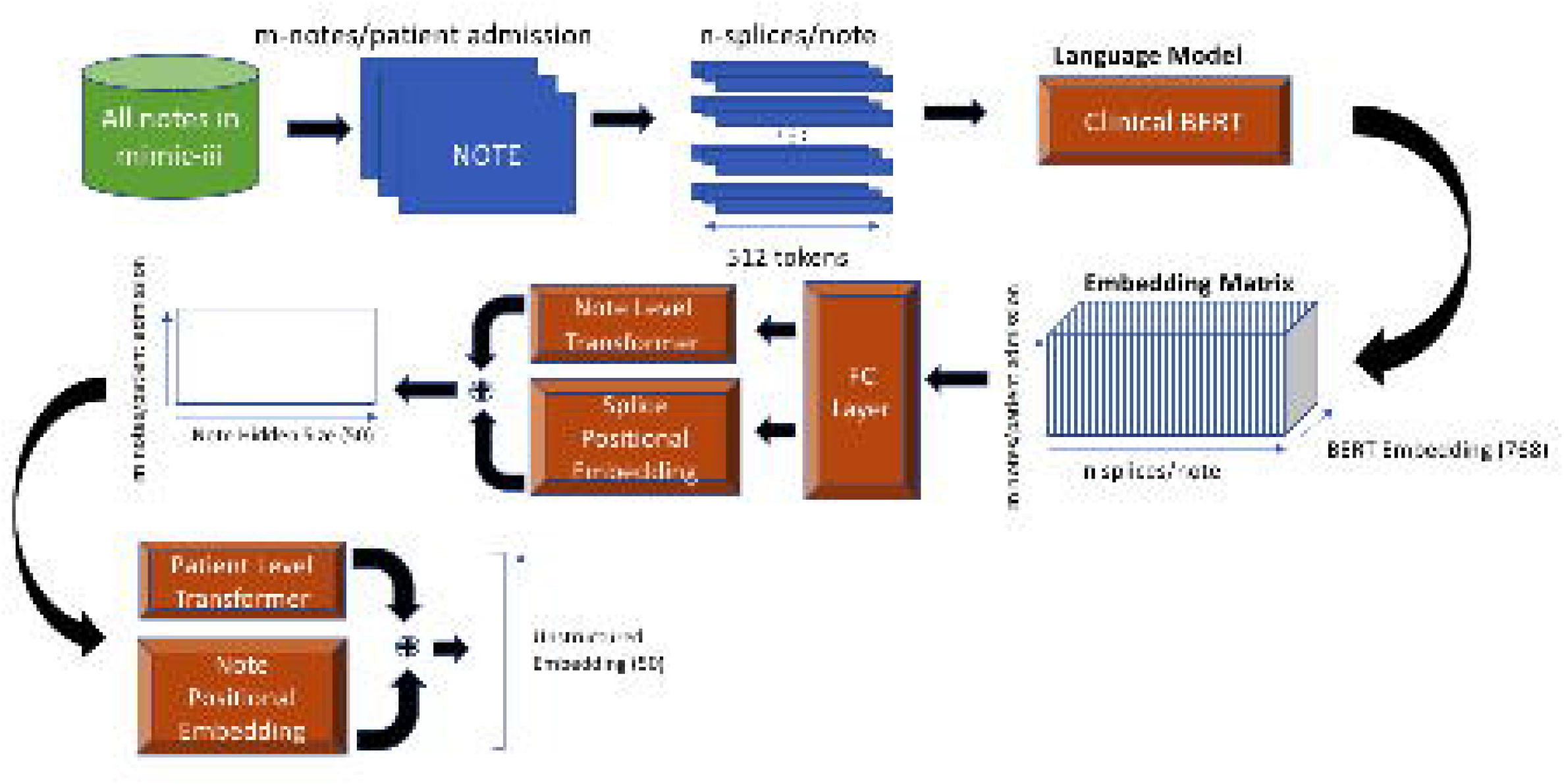
The architecture for a Hierarchical Attention Model using Transformers to process Clinical Notes

**Figure 3:**
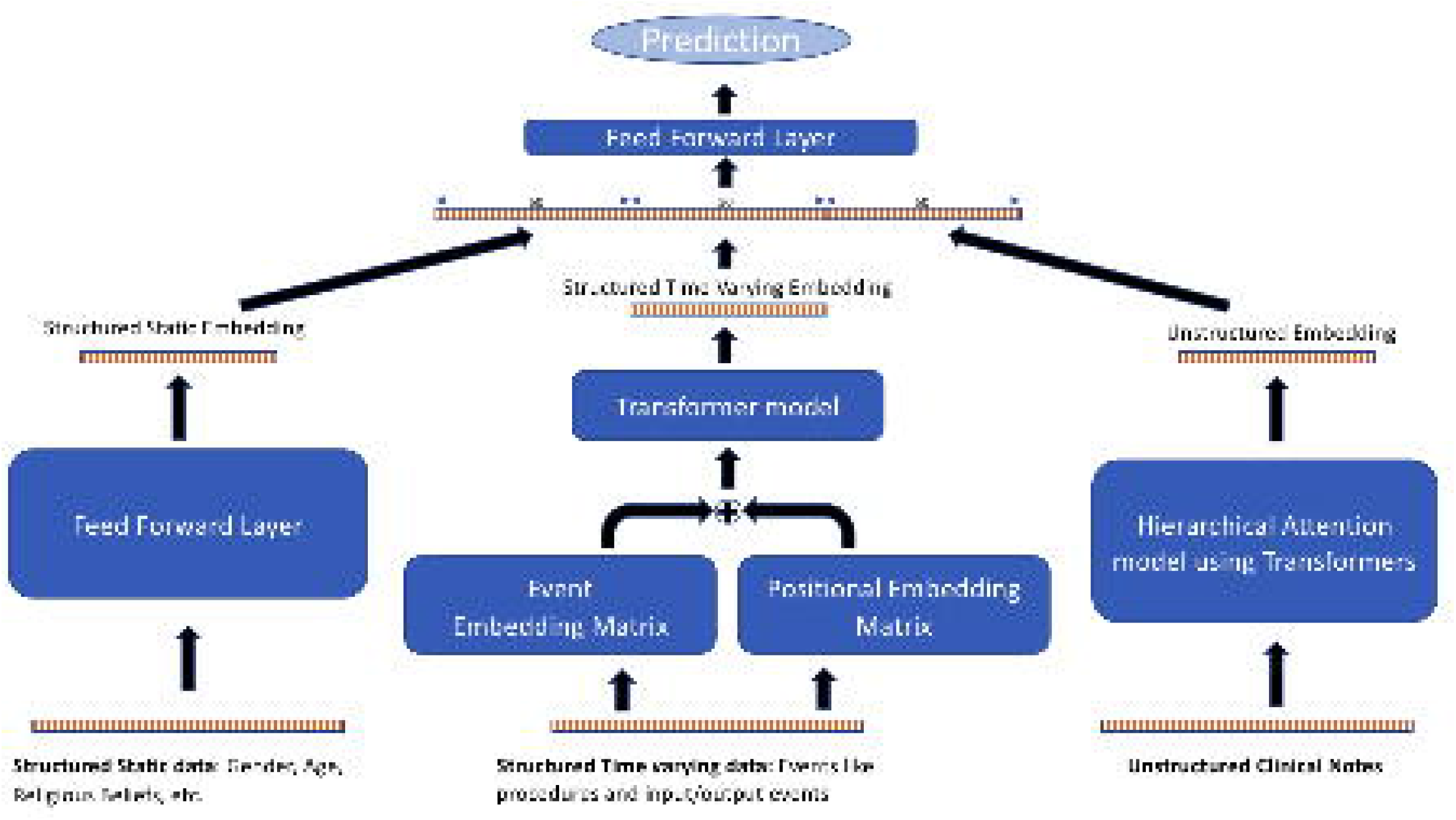
Deep Learning Model architecture that takes in patient EHR data and predicts a binary outcome

1. The structured static data containing categorical features was first converted to indicator variables. A vector of size 45 was obtained for every patient admission, which was then converted to a “structured static embedding” of size 50 by passing it through a feed forward layer.
2. The events in the structured time varying data were first sorted chronologically, following which each event was converted into a one hot encoding. We then obtained a sequence of encoded events for each patient admission. Across all patients, we identified 4,483 unique events with the median number of events per patient being 125. Based off of this mean, we selected a maximum of 150 most recent events for every patient and appended a *start* and *end* token to each sequence. We then passed this sequence of events through two different embedding layers (embedding size of 50). The first embedding layer produced event embeddings while the second embedding layer produced positional embeddings. The positional embeddings were included in the model architecture as we felt relative positions of an event with respect to other events in the sequence was an important signal for predicting the required outcomes. We added the embeddings from these two layers and subsequently fed it into a Transformer layer to obtain contextual event embeddings. The contextual embedding of the *start* token was finally taken as the “structured time varying embedding” of the patient for that specific hospital admission.
3. As each hospital admission of a patient could have multiple clinical notes, we first chronologically sorted the notes and retrieved up to 40 of their most recent notes (just prior to the opioid prescription or OUD diagnoses if a positive example). For the negative examples (patients without an OUD diagnosis or opioid prescription depending on the model), we obtained the last 40 clinical notes. We then obtained note embeddings using a Hierarchical Transformer model as shown in **Figure 2** using the following method. We first tokenize each note using a word piece tokenizer [48], following which we split the tokenized notes into splices of 510 tokens each and append a *[CLS]* and *[SEP]* token to the beginning and end of each splice (the maximum size of the input sequence accepted by a BERT model is 512 [43]). Next, we choose up to 10 note splices per note (which results in a maximum of ∼5100 tokens/note) and obtain embeddings for the respective splices using ClinicalBERT. We reduce the dimension of the splice BERT embeddings to 50 by passing them through a feed forward network. We do this to reduce the memory requirement for the following layers. The reduced BERT embeddings are then passed through two consecutive Transformer Layers after adding positional embeddings. The first Transformer uses attention across splices within a particular note. It calculates a contextualized embedding for the [CLS] token of the first splice which is taken as the note embedding. The second transformer model takes the note embeddings for a maximum of 40 notes calculated by the previous layer and uses attention to calculate a contextualized embedding across all notes. This is finally taken as the “Unstructured Embedding” of the patient admission.
4. The embeddings of the first three components are finally concatenated together and passed through a Feed Forward Layer to obtain a prediction for the binary variable of interest.

We first create a balanced dataset containing an equal number of examples per class for each task (predicting opioid prescription and OUD). Next, we set aside a random subset of the data (10%) as the test set and use the remaining data for training and validating the classification models. Both models were trained for 15 epochs with stochastic gradient descent (learning rate = 0.005 and momentum = 0.9) and Binary Cross Entropy Loss across 10 GPUs with a batch size of 10. As Deep Learning models have non-convex objective functions, they perform slightly differently every time they are trained on the same dataset. As a result, for each of the 2 tasks, we trained a model 10 times and reported the performance metrics on the test sets as the mean ± standard deviation across the 10 models **(Table 2)**.

**Table 2:**
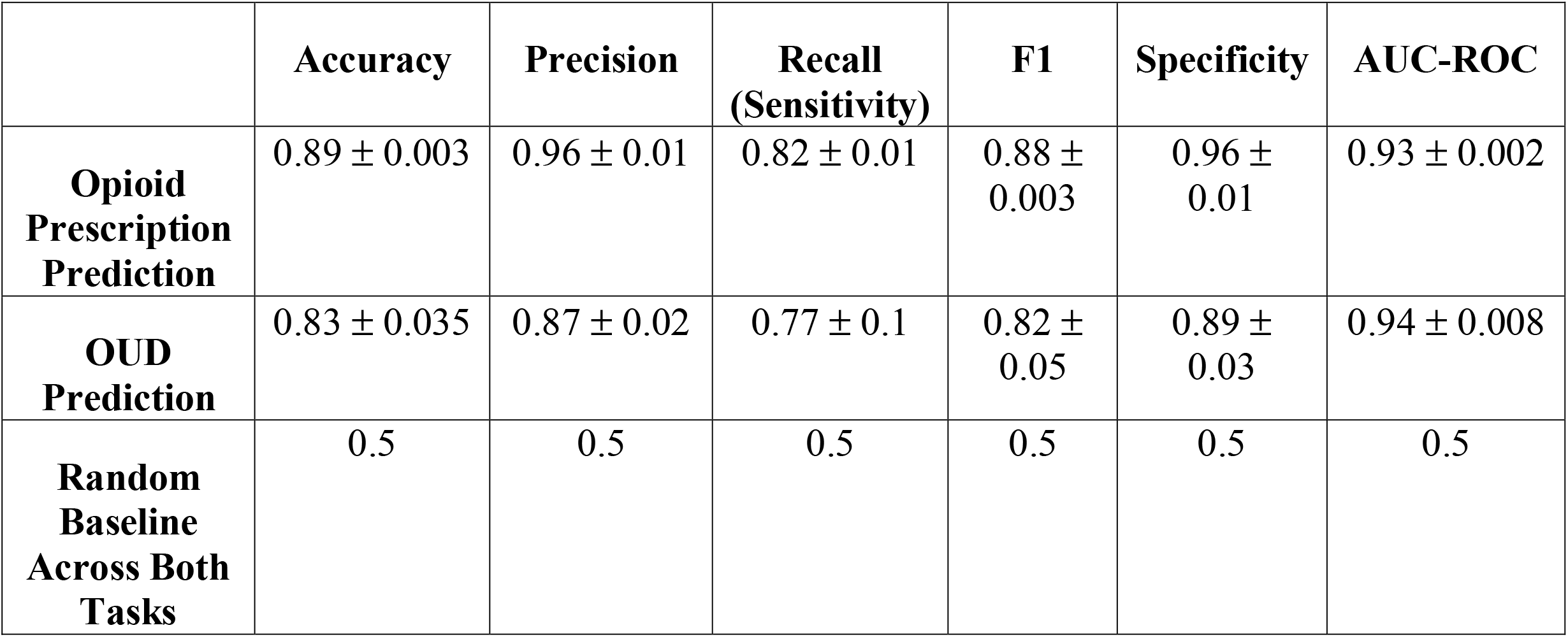
Performance Metrics for the Deep Learning models across the 2 tasks and 10 iterations per task

### 2.3 Model and Data Insights

Since Deep Learning models are black boxes making them hard to explain, we also trained Logistic Regression (LogReg) models across every combination of task (opioid prescription prediction and OUD prediction) and data type (Structured Static Data, Structured Time Varying Data and Unstructured Clinical Notes). Examining the top features considered by the LogReg models could give us some insights into the kind of decisions made by the Deep Learning models in predicting the respective outcomes. The LogReg models were trained on the presence or absence of categorical features in the Structured Static Data **(Table 3)** and the Structured Time Varying Data (**Table 3)**. For the Unstructured Clinical Notes, the text data was first tokenized into unigram, bigram and trigram features, converted to lower-case, and represented as a binary vector (containing a “1” at a particular index if the corresponding ngram was present in the text) that was then used to train a LogReg model (**Table 4**). The 20,000 most frequent ngrams in the clinical notes were considered while constructing these binary vectors.

**Table 3:**
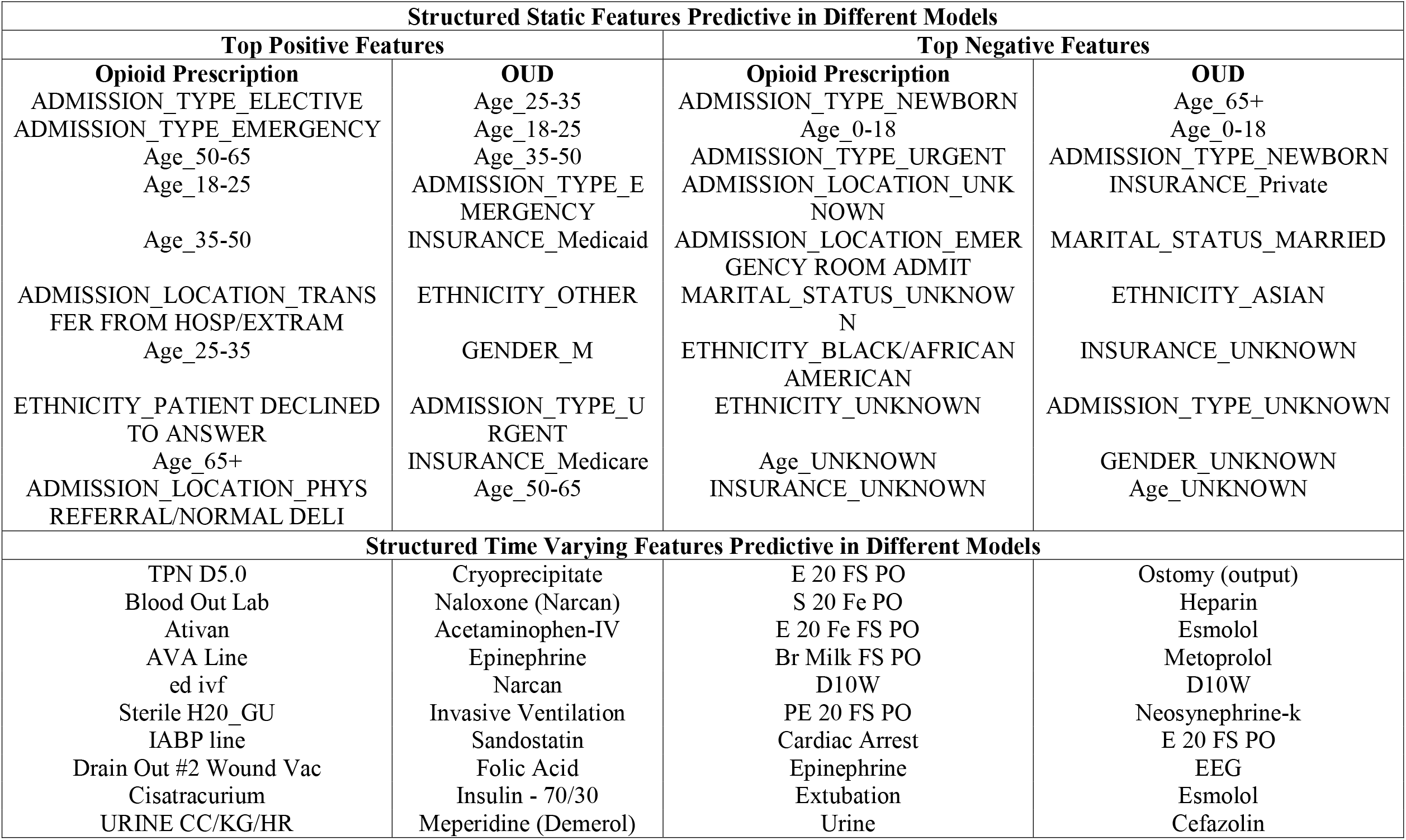
Top 10 Positive and Negative Features considered by the LogReg models across the 2 tasks when considering only Structured Static Features and Structured Time Varying Features respectively

| **Structured Static Features Predictive in Different Models** |  |  |  |
| --- | --- | --- | --- |
| **Top Positive Features** |  | **Top Negative Features** |  |
| **Opioid Prescription**<br>ADMISSION_TYPE_ELECTIVE<br>ADMISSION_TYPE_EMERGENCY<br>Age_50-65<br>Age_18-25<br>Age_35-50<br>ADMISSION_LOCATION_TRANSFER FROM HOSP/EXTRAM<br>Age_25-35<br>ETHNICITY_PATIENT DECLINED TO ANSWER<br>Age_65+<br>ADMISSION_LOCATION_PHYS REFERRAL/NORMAL DELI | **OUD**<br>Age_25-35<br>Age_18-25<br>Age_35-50<br>ADMISSION_TYPE_EMERGENCY<br>INSURANCE_Medicaid<br>ETHNICITY_OTHER<br>GENDER_M<br>ADMISSION_TYPE_URGENT<br>INSURANCE_Medicare<br>Age_50-65 | **Opioid Prescription**<br>ADMISSION_TYPE_NEWBORN<br>Age_0-18<br>ADMISSION_TYPE_URGENT<br>ADMISSION_LOCATION_UNKNOWN<br>ADMISSION_LOCATION_EMERGENCY ROOM ADMIT<br>MARITAL_STATUS_UNKNOWN<br>ETHNICITY_BLACK/AFRICAN AMERICAN<br>ETHNICITY_UNKNOWN<br>Age_UNKNOWN<br>INSURANCE_UNKNOWN | **OUD**<br>Age_65+<br>Age_0-18<br>ADMISSION_TYPE_NEWBORN<br>INSURANCE_Private<br>MARITAL_STATUS_MARRIED<br>ETHNICITY_ASIAN<br>INSURANCE_UNKNOWN<br>ADMISSION_TYPE_UNKNOWN<br>GENDER_UNKNOWN<br>Age_UNKNOWN |
| **Structured Time Varying Features Predictive in Different Models** |  |  |  |
| TPN D5.0<br>Blood Out Lab<br>Ativan<br>AVA Line<br>ed ivf<br>Sterile H2O_GU<br>IABP line<br>Drain Out #2 Wound Vac<br>Cisatracurium<br>URINE CC/KG/HR | Cryoprecipitate<br>Naloxone (Narcan)<br>Acetaminophen-IV<br>Epinephrine<br>Narcan<br>Invasive Ventilation<br>Sandostatin<br>Folic Acid<br>Insulin - 70/30<br>Meperidine (Demerol) | E 20 FS PO<br>S 20 Fe PO<br>E 20 Fe FS PO<br>Br Milk FS PO<br>D10W<br>PE 20 FS PO<br>Cardiac Arrest<br>Epinephrine<br>Extubation<br>Urine | Ostomy (output)<br>Heparin<br>Esmolol<br>Metoprolol<br>D10W<br>Neosynephrine-k<br>E 20 FS PO<br>EEG<br>Esmolol<br>Cefazolin |

**Table 4:**
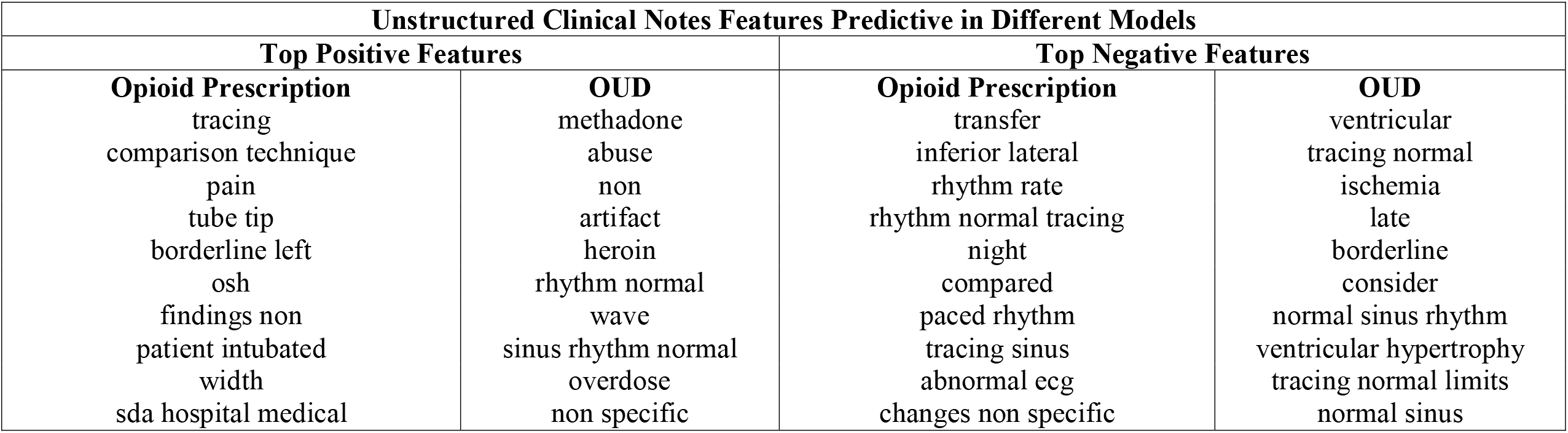
Top 10 Positive and Negative Features considered by the LogReg models across the 2 tasks when considering only Unstructured Clinical Notes

## 3. RESULTS

### 3.1 Dataset

Using the techniques mentioned in the previous sections, we identified 37,237 unique admissions (HADM_IDs) of patients who were prescribed at least one opioid during their hospital admission. Similarly, we identified 763 unique admissions (HADM_IDs) of patients who were diagnosed with OUD. Additionally, the distribution of patient demographics across the positive and negative examples for both tasks is shown in **Table 1**.

### 3.2 Prediction Models

Details for the size of the training, validation and test sets across both the tasks are shown in **Table 3** of the Supplement. The performance metrics of the models on the held-out test sets are shown in **Table 2**. To put these results into perspective, a random baseline accuracy for both models is 0.5. Our opioid prescription deep learning prediction algorithm performed well with an F1 score of 0.88 +/- 0.003 and an AUC-ROC of 0.93 +/- 0.002. The OUD deep learning prediction algorithm also performed well with an F1 score of 0.82 +/- 0.05 (slightly lower than our opioid prescription algorithm) and an AUCROC was 0.94 +/- 0.008 (slightly higher than our opioid prescription algorithm). This indicates that deep learning can be used to accurately predict both opioid prescription behavior and OUD diagnosis.

### 3.3 Important Features

The top positive features of each LogReg model across the 3 different data types and the 2 tasks are shown in **Table 3** and **Table 4**, and provide some insights into the kind of patient features that the deep learning models might look for while making predictions. For example, admission types and age groups were informative at either predicting opioid prescription or non-prescribing (negative features) along with OUD diagnosis or non-diagnosis (**Table 3)**. Newborn admissions were less likely to be diagnosed with either OUD or prescribed an opioid while emergency admissions were more likely to be diagnosed with an OUD or prescribed an opioid (**Table 3**). Similarly, elective admission types were more likely to be prescribed an opioid, but no relationship was found with OUD diagnosis (**Table 3**).

## 4. DISCUSSION

### 4.1 Prediction of Opioid Prescribing

Scanning through the literature, we were unable to find past research that focused on predicting likelihood of opioid prescriptions based on EHRs. This work therefore makes an important contribution of filling this gap by showing that such high performing models can be built with an F1 score of 0.88 ± 0.003 and an AUC-ROC score of 0.93 ± 0.002 (**Table 2**).

### 4.2 Prediction of Opioid Use Disorder (OUD)

We first compare the performance of our deep learning model to past research. SOAPP-R [49] is a self-report questionnaire used to determine which patients are high-risk for opioid misuse. It is one of the most validated tools for opioid dependence identification [35], and based on literature there is some variation in sensitivity and specificity. The sensitivity of SOAPP was reported to range from 67% to 81% [50][51][52] and the specificity ranged from 52% to 68%[50][51][52]. In our OUD model, we achieved a sensitivity score of 0.77 ± 0.1 and specificity score of 0.89 ±0.03 **(Table 2)**. Therefore, our model outperformed SOAPP in terms of specificity and achieved equivalent sensitivity.

In other research [36], a machine learning model was built on EHRs to predict opioid dependence, which obtained a mean area under the receiver operating characteristic curve (AUC-ROC) of 92%. An LSTM model with attention built in [37] to predict overdose risk achieved an F1 score of 0.78 and an AUC-ROC of 0.8449. The OUD model built in this paper achieved an F1 score of 0.82 ± 0.05, and an AUC-ROC score of 0.94 ± 0.008 **(Table 2)**. Although, we outperformed the prior work in this area in terms of AUC-ROC and F1 score, it is important to note that opioid dependence studies carried out often have variations in study periods, sample sizes, definitions, types of databases, and structured documentations, and therefore some of the differences in model performances built across these studies may make the models not be directly comparable.

Authors in [35] conclude that age and gender are the most consistent demographic variables in predicting opioid abuse. The features for OUD in **Table 3** show similar findings in our experiments, with the top three positive features being age groups, and male gender being positively correlated with OUD. These researchers also noted that certain medication variables were among the most indicative features that occurred across papers that they reviewed [35]. We obtained similar results with medications such as naloxone, acetaminophen-IV and epinephrine **(Table 3)**, and methadone and heroin (**Table 4)** being predictive features for OUD. This is also similar to the findings in [37] where the authors mention that the medication class of opioids are among the most important features for OUD prediction. Similar to [36], we found methadone (**Table 4)** and folic acid (**Table 3)** to be present in EHRs of OUD patients.

### 4.3 Our Deep Learning Models Overcome Known Limitations in the Field

Our models overcome some limitations that researchers studying opioid dependence in EHR data have faced in the past. A lot of previous research relies on domain knowledge from clinical experts for feature engineering, as mentioned in [37]. This typically requires a lot of effort and can be quite cumbersome and expensive. With our Deep Learning models, features are learned automatically as part of the training process, providing an inexpensive way of identifying such medical outcomes of interest. Researchers also often construct predictive models on structured parts of the EHRs, often ignoring the unstructured part which can account for 80% of the total health data [35]. In this paper, we build models that take in information present in both the structured and unstructured parts of the EHRs to make informed decisions by using state-of-the-art models from NLP. Researchers in [40] attempt to use unstructured clinical text to predict opioid related overdoses. However, their approach is expensive as it relies on domain experts to manually develop “rule trees”.

### 4.4 Limitations and Future Work

The approaches mentioned in this paper have some limitations. Researchers in [35] mention that ICD codes can often understate the actual number of patients exhibiting the target categorization [53, 54] due to an intentional lack of documentation, or in other cases, prescribers trying to avoid stigmatizing patients with opioid problems. As a result, some patients who have an opioid problem may be overlooked. This could result in some False Negatives in our training data hurting model generalizability. Additionally, we only report model performance metrics on a test-set obtained from the same data distribution as the training data. This may only be partly indicative of the model’s performance in a general setting. Ideally, we would also want to evaluate the model on an out-of-distribution test dataset, but due to the difficulty in obtaining another dataset for evaluation, this was not possible for this study but remains a subject of future work. Lastly, due to population variations and procedural differences between the clinic in which this tool was created and the target clinic where this tool might be adopted, there could be some barriers to its adoption.

## 5. CONCLUSION

In this paper, we have developed an informatics algorithm that trains two deep learning models over patient EHRs to predict how likely they are of being prescribed an opioid and developing OUD. We utilize both the structured and unstructured parts of the EHR and show that it is possible to predict these outcomes with an accuracy of 0.89 ± 0.003 and 0.83 ± 0.035 respectively. As the opioid epidemic continues to grow in the United States, utilizing such models in the patient treatment process will help healthcare workers bring the situation under control.

## Data Availability

This paper uses the MIMIC-III database that is publicly available at https://physionet.org/content/mimiciii/1.4/ provided users due requisite training and obtain Institutional Review Board approval for their project.

