## Supplement for "A Deep Learning Method to Detect Opioid Prescription and Opioid Use Disorder from Electronic Health Records"

### Table of Contents

*Tables*

Table 1: List of Opioids used in obtaining positive training examples for the Opioid Prescription Prediction Model……..............................................................................................................…2

Table 2: A list of ICD9 codes used to identify OUD patients………………………………..........3

Table 3: Train, Validation and Test Set sizes for the 2 tasks…………………………………………………………………………………….......4

*Table 1: List of Opioids used in obtaining positive training examples for the Opioid Prescription Prediction Model*

| **Opioid Medication Name** |
|---|
| Codeine |
| Hydromorphone |
| MS_Contin |
| Percocet |
| Ultracet |
| Guiatuss_AC |
| Dilaudid |
| Lorcet |
| Nalbuphine |
| Roxicet |
| Ultram |
| Robitussin_DM |
| Duragesic |
| Lortab |
| Norco |
| Roxicodone |
| Vicodin |
| Safetussin_DM |
| Endocet |
| Meperidine |
| Oxycodone |
| Tramadol |
| Delsym |
| Tussionex |
| Fentanyl |
| Morphine |
| Oxycontin |
| Tylenol_with_Codeine |
| Dextromethorphan |
| Wal_Tussin_Cough |

*Table 2: A list of ICD9 codes used to identify OUD patients*

| ICD9 Code | Description |
|---|---|
| 30400 | Opioid type dependence, unspecified |
| 30401 | Opioid type dependence, continuous |
| 30402 | Opioid type dependence, episodic |
| 30403 | Opioid type dependence, in remission |
| 30470 | Combinations of opioid type drug with any other drug dependence, unspecified |
| 30471 | Combinations of opioid type drug with any other drug dependence, continuous |
| 30472 | Combinations of opioid type drug with any other drug dependence, episodic |
| 30473 | Combinations of opioid type drug with any other drug dependence, in remission |
| 30550 | Opioid abuse, unspecified |
| 30551 | Opioid abuse, continuous |
| 30552 | Opioid abuse, episodic |
| 30553 | Opioid abuse, in remission |

*Table 3: Train, Validation and Test Set sizes for the 2 tasks*

| | **Opioid Prescription Prediction** | | **OUD Prediction** | |
|---|---|---|---|---|
| | **# Positive** | **# Negative** | **# Positive** | **# Negative** |
| **Train** | 17669 | 17669 | 689 | 689 |
| **Validation** | 929 | 929 | 72 | 72 |
| **Test** | 1895 | 1895 | 76 | 76 |
